# Genome-wide trans-ethnic meta-analysis identifies novel susceptibility loci for childhood acute lymphoblastic leukemia

**DOI:** 10.1101/2021.05.07.21256849

**Authors:** Soyoung Jeon, Adam J. de Smith, Shaobo Li, Minhui Chen, Tsz Fung Chan, Ivo S. Muskens, Libby M. Morimoto, Andrew T. DeWan, Nicholas Mancuso, Catherine Metayer, Xiaomei Ma, Joseph L. Wiemels, Charleston W.K. Chiang

**Affiliations:** Center for Genetic Epidemiology, Department of Preventive Medicine, Keck School of Medicine, University of Southern California, Los Angeles, CA; Cancer Biology and Genomics Graduate Program, Program in Biological and Biomedical Sciences, Keck School of Medicine, University of Southern California, Los Angeles, CA; Division of Epidemiology & Biostatistics, School of Public Health, University of California, Berkeley, CA; Center for Perinatal, Pediatric and Environmental Epidemiology, Yale School of Public Health, New Haven, CT; Department of Chronic Disease Epidemiology, Yale School of Public Health, New Haven, CT; Department of Quantitative and Computational Biology, University of Southern California, Los Angeles, CA; Norris Comprehensive Cancer Center, Keck School of Medicine, University of Southern California, Los Angeles, CA

## Abstract

The risk of childhood acute lymphoblastic leukemia (ALL) differs across ethnic groups and there exist gaps in our understanding of the genetic risk of ALL as most studies took place in populations of predominantly European ancestries. In an effort to address these limitations, we performed a genetic meta-analysis of ALL in 76,317 participants across four ethnic groups, including 17,814 non-European individuals and 3,482 total cases. We replicated 15 out of 16 previously identified loci associated with ALL in our trans-ethnic analysis. We further identified five novel associations at genome-wide significance, including three novel loci and two secondary associations at previously known loci (17q12 and near *CEBPE*). The three putatively novel loci (rs9376090 near *MYB/HBS1L*, rs10998283 near *TET1*, and rs9415680 near *JMJD1C/NRBF2*) were previously shown to be associated with multiple blood cell traits and other hematopoietic cancers. Polygenic risk scores constructed from our trans-ethnic meta-analysis showed similar efficacy in independent Latino (LAT) and non-Latino white (NLW) ALL cohorts (AUC ∼ 0.67-0.68) and could partly explain the increased risk of ALL in LAT compared to NLW. Cross-population analysis also showed high but significantly less than 100% genetic correlation between LAT and NLW, suggesting potential differences in the underlying genetic architecture between ethnic groups. In summary, our findings enhance the understanding of genetic contribution to ALL risk across diverse populations and highlight the importance to include multiple ethnic groups in GWAS.

**Key Points:**

- We identified three novel loci on childhood acute lymphoblastic leukemia near genes previously associated with multiple blood cell traits
- Polygenic risk scores using known and novel risk variants showed similar efficacy in Latino and non-Latino white Americans

## Introduction

Acute lymphoblastic leukemia (ALL) is the most common type of childhood cancer worldwide, with substantial racial and ethnic differences in incidence and treatment outcome^1,2^. Previous genome-wide association studies (GWAS) have confirmed the genetic basis of ALL susceptibility by identifying a number of risk loci for childhood ALL^3,4,5(p12),6–8^ and estimating the heritability to be 21% (ref.^9^). However, the known risk loci together account for a relatively small portion of the total variance in genetic risk of ALL^9^, suggesting that additional susceptibility alleles may be discovered in larger studies. Furthermore, these studies were generally performed in cohorts with a predominantly European ancestry. Latino children have the highest risk of ALL in the United States, with an incidence rate ∼15-40% higher than in non-Latino whites^10–12^ and an increased chance of relapse and poorer overall survival^13,14^. Yet, we have a limited understanding of the genetic architecture of ALL in non-European populations and the generalizability of findings from existing GWAS to non-European populations (but see recent efforts for studying the genetic etiology of childhood ALL in Latinos^15–17^). While environmental or social factors likely underlie some if not the majority of the differences in risk between ethnic groups, there may also be a difference in the genetic risk architecture that modulates risk across ethnic groups and would argue for the greater inclusion of other ethnic groups in genetic studies of ALL.

Given this context, we performed a trans-ethnic GWAS of childhood ALL in a discovery panel consisting of 76,317 individuals from an assembled multi-ethnic cohort. We note the complexity of discussing race, ethnicity and ancestry in a genetic study. As a convention, we used the following terms and abbreviations to refer to each ethnic group in our study: African American (AFR), East Asian (EAS), Latino American (LAT), and non-Latino white (NLW). These population labels are largely based on self-reported ethnic identity and we confirmed that they largely correlate with genetic ancestry as defined by the reference populations in 1000 Genomes^18^ (Methods). Our cohort consisted of 3,482 cases and 72,835 controls for an effective sample size of 13,292, which is, to our knowledge, the largest trans-ethnic GWAS for ALL to date. We identified three novel ALL risk loci and tested the novel findings from our discovery panel in two additional independent cohorts. We further compared the efficacy of polygenic risk scores (PRS) to stratify individuals based on their risk of ALL in the two largest subgroups of our data, LAT and NLW. PRS models are known to be poorly transferred to non-European populations^19^, but multi-ethnic designs may be more effective in identifying alleles with shared effects across population without explicit fine-mapping and produce more comparable PRS models between populations^20,21^. Finally, we leveraged our genome-wide summary statistics to contrast the genetic architecture of ALL between LAT and NLW populations.

## Materials and methods

### Study Cohorts

The California Childhood Cancer Record Linkage Project (CCRLP) includes all children born in California during 1982-2009 and diagnosed with ALL at the age of 0-14 years per California Cancer Registry records. Children who were born in California during the same period and not reported to California Cancer Registry as having any childhood cancer were considered potential controls. Detailed information on sample matching, preparation and genotyping has been previously described.^4^ Because ALL is a rare childhood cancer, for the purpose of a genetic study we followed previous practice^4^ and incorporated additional controls using adult individuals from the Kaiser Resource for Genetic Epidemiology Research on Aging Cohort (GERA; dbGaP accession: phs000788.v1.p2). The GERA cohort was chosen because a very similar genotyping platform had been used: Affymetrix Axiom World arrays. For replications we included two independent ALL cohorts: (1) individuals of predominantly European ancestry from the Children’s Oncology Group (COG; dbGAP accession: phs000638.v1.p1) as cases and from Wellcome Trust Case–Control Consortium^22^ (WTCCC) as controls; and (2) individuals of European and Latino ancestry from the California Childhood Leukemia Study (CCLS), a non-overlapping California case-control study (1995-2008).^23^ The quality control and imputation for both the discovery and replication cohorts were conducted in ethnic strata and generally followed previous pipelines of ALL GWAS, but with additional attention paid to incorporate the entire GERA cohort and ensuring data quality post-imputation. See Supplemental Methods for details. This study was approved by Institutional Review Boards at the California Health and Human Services Agency, University of Southern California, Yale University, and the University of California San Francisco.

### Association Testing

We used SNPTEST^24^ (v2.5.2) to test the association between imputed genotype dosage and case-control status in logistic regression, after adjusting for the top 20 principal components (PCs). Sex was not included as a covariate, and we found sex was not correlated with genotype dosage of any of the putatively associated SNPs (data not shown). Results from the four ethnic-stratified analyses were combined via the fixed-effect meta-analysis with variance weighting using METAL^25^. Only variants passing QC in at least three of the four ethnic groups were meta-analyzed. A genome-wide threshold of 5 × 10^−8^ was used for significance in the discovery stage. A Bonferroni-corrected significance of 0.00312 (=0.05/16) was used for replication of previously reported susceptibility variants^3–8,26–28^. Cochran’s *Q*-test for heterogeneity was performed using METAL^25^. To perform conditional analysis in identifying secondary associations within a locus, the lead SNP was additionally included in the regression model, again using 5 × 10^−8^ as threshold for significance.

### Polygenic Risk Score Analysis

Polygenic risk scores (PRS) for ALL were constructed using PLINK (v2.0) by summing the genotype dosages of risk alleles, each weighted by its effect size from our discovery GWAS meta-analysis. PRS were constructed based on: (1) lead SNPs in the 16 known loci (N = 18 SNPs, including variants from the two secondary signals in *IKZF1* and *CDKN2A/B* that were previously reported; for which we used the corresponding effect sizes from conditional analysis), and (2) by additionally including the novel hits (N = 23 SNPs, including the additional 3 novel loci and 2 novel conditional associations). Associations between PRS and case-control status for ALL were tested in each group adjusting for 20 PCs using R. To evaluate the predictive power of PRS, Area Under the receiver operating characteristic Curve (AUC) were calculated using pROC package^29^ in R.

### Genetic architecture of ALL within and between populations

To investigate the genetic architecture of ALL and contrasting this architecture between NLW and LAT populations, we estimated the percentage of familial relative risk (FRR) explained by associated variants individually or in aggregate, the heritability ascribable to all post-QC imputed SNPs with MAF ≥ 0.05, the genetic correlation between NLW and LAT, and the genome-wide proportion of causal variants that are population-specific or population-shared. See the Supplemental Methods for details.

## Results

### Trans-ethnic Genetic Associations with ALL

We performed a trans-ethnic meta-analysis GWAS for childhood ALL. After quality control filtering, our dataset consisted of 3,482 cases and 72,835 controls (Supplementary Table S1; Methods) in total. In contrast to the previous trans-ethnic analysis^4^, we included additional controls for NLW and added the EAS cohort. Furthermore, we tested the association at 7,628,894 imputed SNPs, including low frequency (MAF between 1-5%) variants that were not previously systematically tested. We aggregated the summary statistics across the four ethnic groups in a fixed-effect meta-analysis. The genomic control inflation factor was 1.022 after excluding 16 previously known ALL-associated loci (Table 1), suggesting our meta-analysis was reasonably robust to any confounding due to population stratification (Figure 1). In total, twelve loci reached genome-wide significance (i.e, P < 5.0 X 10^−8^) in our analysis.

**Table 1.**
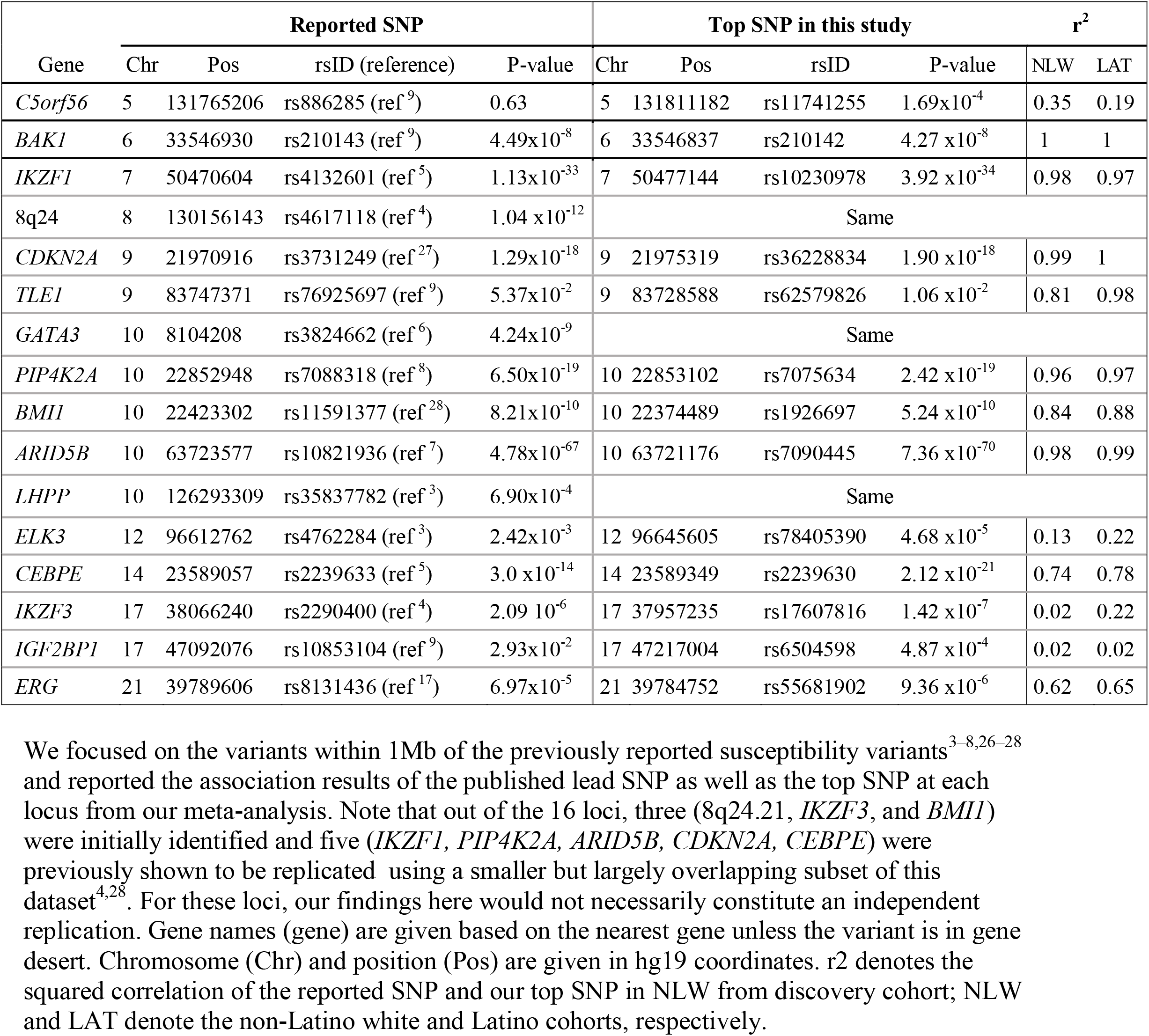
Summary statistics for the reported variants, the top variant in the loci from our meta-analysis, and the linkage disequilibrium between the two variants in NLW and LAT.

**Figure1.**
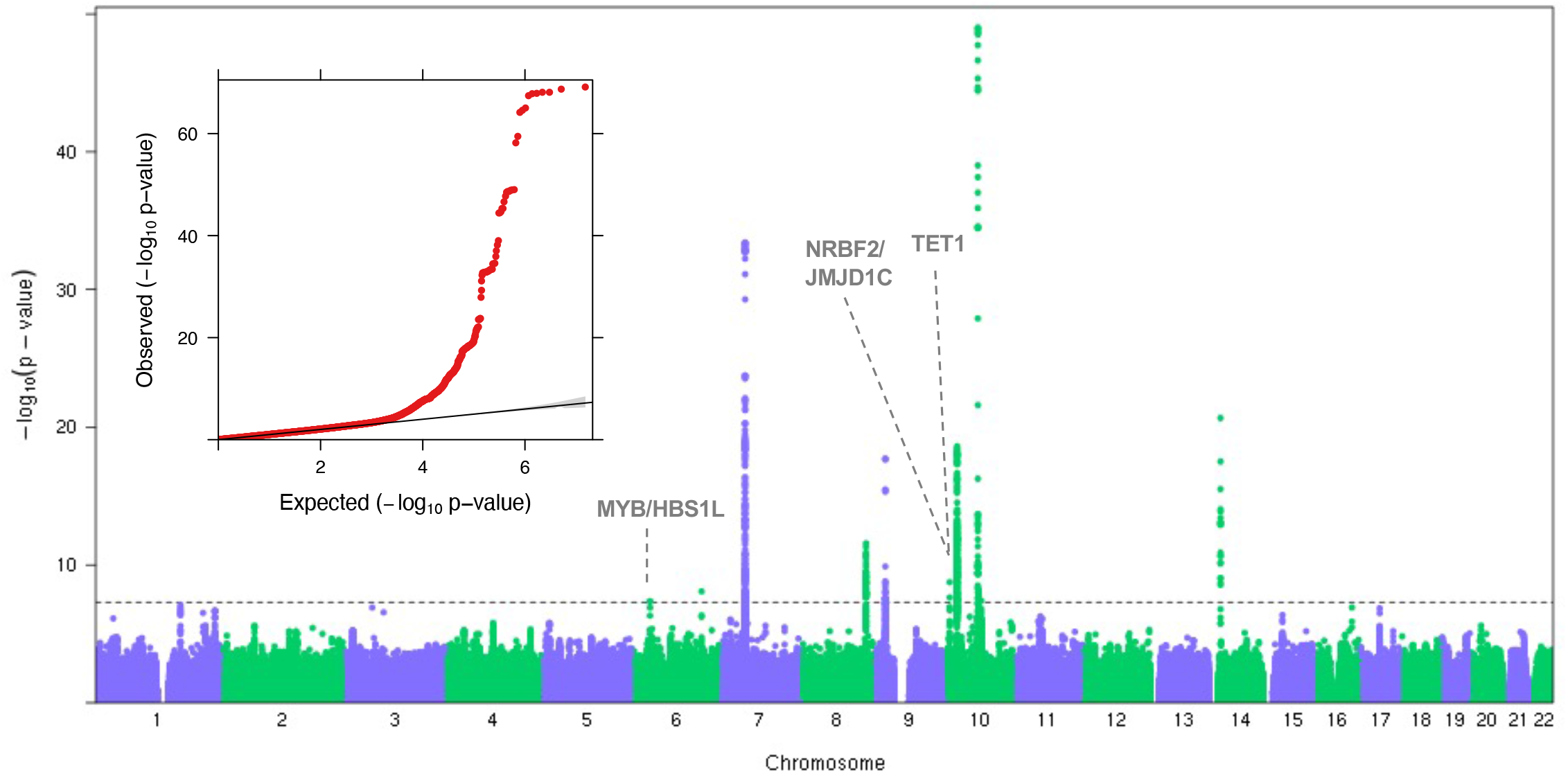
Summary result of the trans-ethnic meta-analysis on ALL. Results of the meta-analysis is represented by the Manhattan plot. The novel loci from this study are marked with dotted lines and labeled with the nearest genes. Significance threshold at genome-wide significance level (5×10^−8^) is marked with a horizontal dashed grey line in the Manhattan plot. The y-axis is truncated at –log10(1×10^−50^) to improve readability. The insert shows the Quantile-Quantile plot. Deviation from the expected p-value distribution is evident only in the tail. There is little evidence of inflation of the test statistics in general as the genomic inflation factor is 1.024.

We found that for the 16 previously published risk loci for ALL^3–9,17,27,28^, all were associated with ALL at the nominal level (P < 0.05) or have a SNP nearby with strong association (Table 1). Nearly all of the published risk SNPs show consistent direction of effects across ethnic groups (13/16 SNPs with heterogeneity P-value > 0.05; P = 0.0384, 0.006, 0, 0.000259 for AFR, EAS, LAT, NLW respectively for consistent direction of effect by the sign test, Supplemental Table S2). In some cases, the published SNP is not the SNP with the most significant association in our dataset, though usually our top SNP in the locus is in strong LD with the reported SNP (Table 1). Given the larger sample size and trans-ethnic analysis, the best associated variants in our analysis may reflect the more likely causal / shared association across populations. Two loci at *C5orf56* and *TLE1* are noted. At the *C5orf56* locus on 5q31, the variant previously reported in an independent European-ancestry cohort (rs886285) to be associated with a particular subtype of ALL (HD-ALL)^9^ was not nominally associated with ALL overall (P = 0.63) in our dataset. A weakly linked SNP (rs11741255; r2 = 0.35 in NLW, 0.19 in LAT) in the same locus approximately 20kb away was significantly associated with ALL in our data (P = 1.69×10^−4^) but may reflect a chance association. At the *TLE1* locus on 5q21, neither the published variant nor our top variant in the locus would be considered significantly associated after Bonferroni correction (minimum P = 1.06×10^−2^ for rs62579826), possibly due to heterogeneity driven by EAS in which both the published variant and our top variant are monomorphic^30^.

More importantly, we discovered three putatively novel susceptibility loci: one at 6q23 and two at 10q21 (Figure 1). The strongest association signal in 6q23 is at rs9376090 (P= 8.23 X 10^−9^, OR=1.27) in the intergenic region between *MYB* and *HBS1L* (Figure 2A). This association is mainly driven by NLW presumably due to its large sample size (Supplementary Table S1). In 10q21, there were two independent signals that showed genome-wide significance. One locus was identified with the lead SNP rs9415680 (P=7.27 X 10^−8^, OR=1.20), within a broad association peak, with apparently long-range LD with SNPs covering *NRBF2, JMJD1C*, and parts of *REEP3* (Figure 2B). The second locus in 10q21 was identified 5Mb away, with lead SNP rs10998283 (P=3.92×10^−8^, OR=1.15) in an intronic region in *TET1* (Figure 2C). The association signals for both loci in 10q21 were largely driven by LAT. We used the convention of the nearest genes to refer to these loci for the remainder of the manuscript, acknowledging that they may not be the causal genes.

**Figure2.**
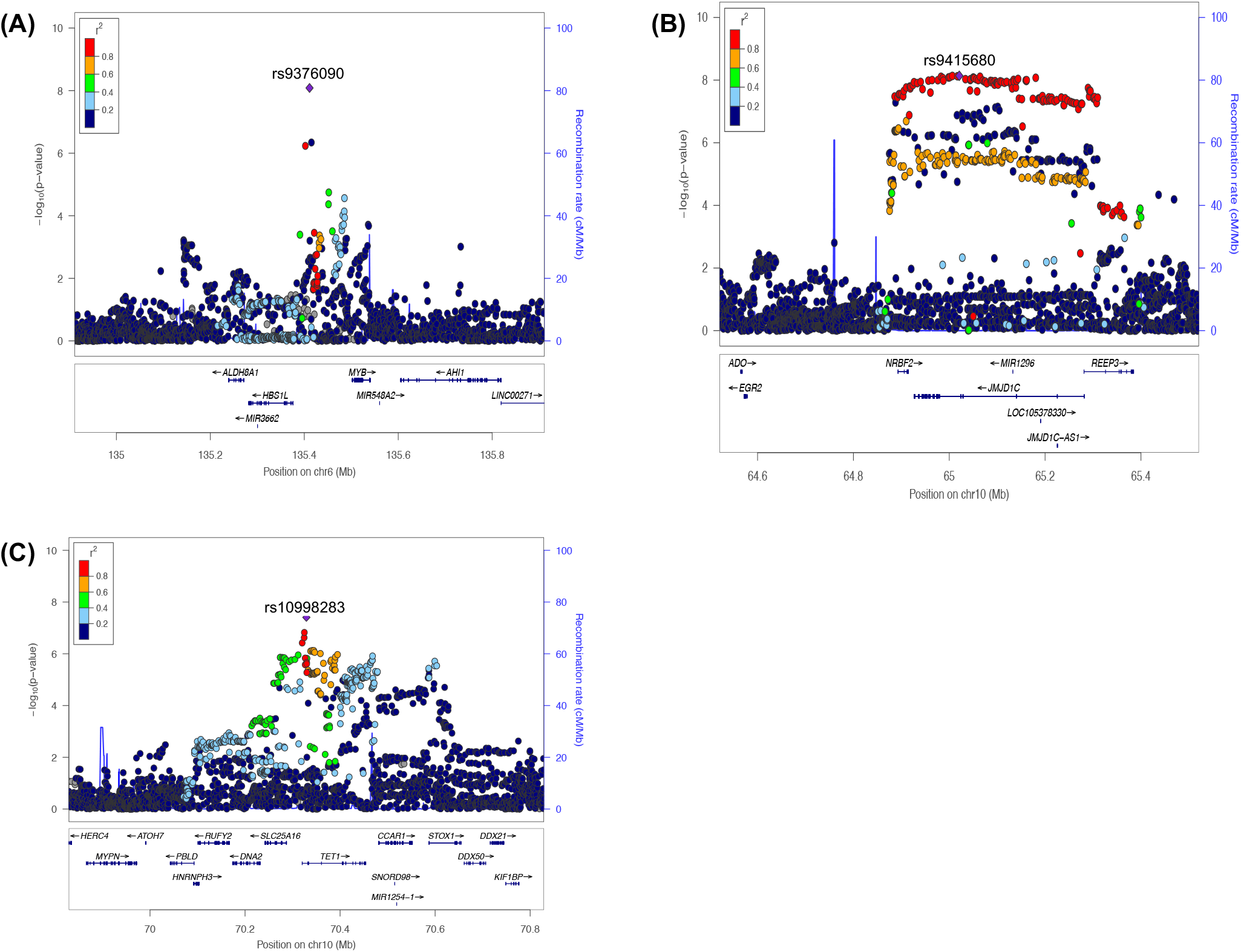
Novel loci associated with childhood ALL in trans-ethnic meta-analysis. LocusZoom plots showing 1 Mb region around the identified loci near (A) MYB/HBS1L on chr6, (B) NRBF2/JMJD1C on chr10, and (C) TET1 on chr10 are shown. Diamond symbol indicates the lead SNP in each locus. Color of remaining SNPs is based on linkage disequilibrium (LD) as measured by r2 with the lead SNP in non-Latino white. All coordinates in x-axis are in hg19.

To replicate our findings in independent datasets, we tested the associations of the three novel variants and their LD proxies (with P < 5 × 10^−7^;n=141) in independent samples from the COG/WTCCC and CCLS cohorts (Methods). For the *MYB/HBS1L* locus, which was driven by NLW in the discovery cohort, we replicated the signal in COG/WTCCC cohort (rs9376090, P_COG_= 4.87×10^−3^, P_COG+discovery analysis_ = 1.23×10^−10^; Supplementary Table S3), but did not replicate in CCLS likely owing to the small sample size of NLW. For the *TET1* locus, in which the original association was driven by LAT in the discovery, three of the four SNPs with P < 5×10^−7^ in the discovery cohort nominally replicated in CCLS. The lead SNP after meta-analyzing the discovery cohort and the replication cohort of CCLS was rs79226025 (P_CCLS_ = 3.04 ×10^−2^, P_CCLS+discovery_ = 6.81 ×10^−9^; Supplementary Table S3). For the *NRBF2* / *JMJD1C* locus, we did not observe an association in the replication cohorts.

We also performed conditional analyses adjusting for the lead SNP at each locus and identified a secondary signal in four out of the 16 previously known loci (Table 2, Figure 3). In all cases, the LD between the secondary hit and the top hit in the locus are low (Table 2). The additional second associations in *CDKN2A* and *IZKF1* loci were previously noted^9(p1)^. In *CEBPE* (rs60820638, P=5.38 ×10^−8^) and 17q12 (rs12944882, P=7.71 ×10^−10^), these secondary signals represent novel associations. In particular, at the *CEBPE* locus, previous reports suggest multiple correlated variants with functional evidence^31,32^. Our analysis is consistent with the two previous variants (rs2239635 and rs2239630) being or tagging the same underlying signal, while the new association we identified (rs60820638) is an independent association (Supplementary Table S4).

**Table 2.** Summary of conditional analysis to identify secondary associations at known loci.

| Gene | Chr | Pos | rsID | Risk allele | OR | P<sub>conditional</sub> | P<sub>discovery</sub> | r<sup>2</sup> |
| --- | --- | --- | --- | --- | --- | --- | --- | --- |
| *IKZF1* | 7 | 50459043 | rs78396808 | A | 1.632 | 3.46x10<sup>-26</sup> | 2.7x10<sup>-16</sup> | *0.06 |
| *CDKN2A/B* | 9 | 21993964 | rs2811711 | T | 1.355 | 7.2x10<sup>-10</sup> | 1.85x10<sup>-11</sup> | 0.01 |
| *CEBPE* | 14 | 23592617 | rs60820638 | A | 1.193 | 5.38x10<sup>-8</sup> | 0.102 | 0.16 |
| *IKZF3* | 17 | 37983492 | rs12944882 | T | 1.204 | 7.71x10<sup>-10</sup> | 2.81x10<sup>-7</sup> | 0.02 |

For each of the four significant association after conditional analysis, we show the genomic coordinates in hg19, effect size (OR), the P-values with or without conditioning on the lead SNP from the discovery meta-analysis in the locus, and the r<sup>2</sup> between the lead SNP and secondary association.
Chr., chromosome; Pos., Position in hg19; OR: Effect size; P<sub>conditional</sub>: p-value from the conditional analysis; P<sub>discovery</sub>: p –value from meta-analysis without conditioning on any SNP; r<sup>2</sup>: squared correlation of the conditioned SNP and the most significantly associated SNP from conditional analysis.
\*calculated in Latino population as the variant was filtered out for low MAF in NLW cohort.

**Figure 3.**
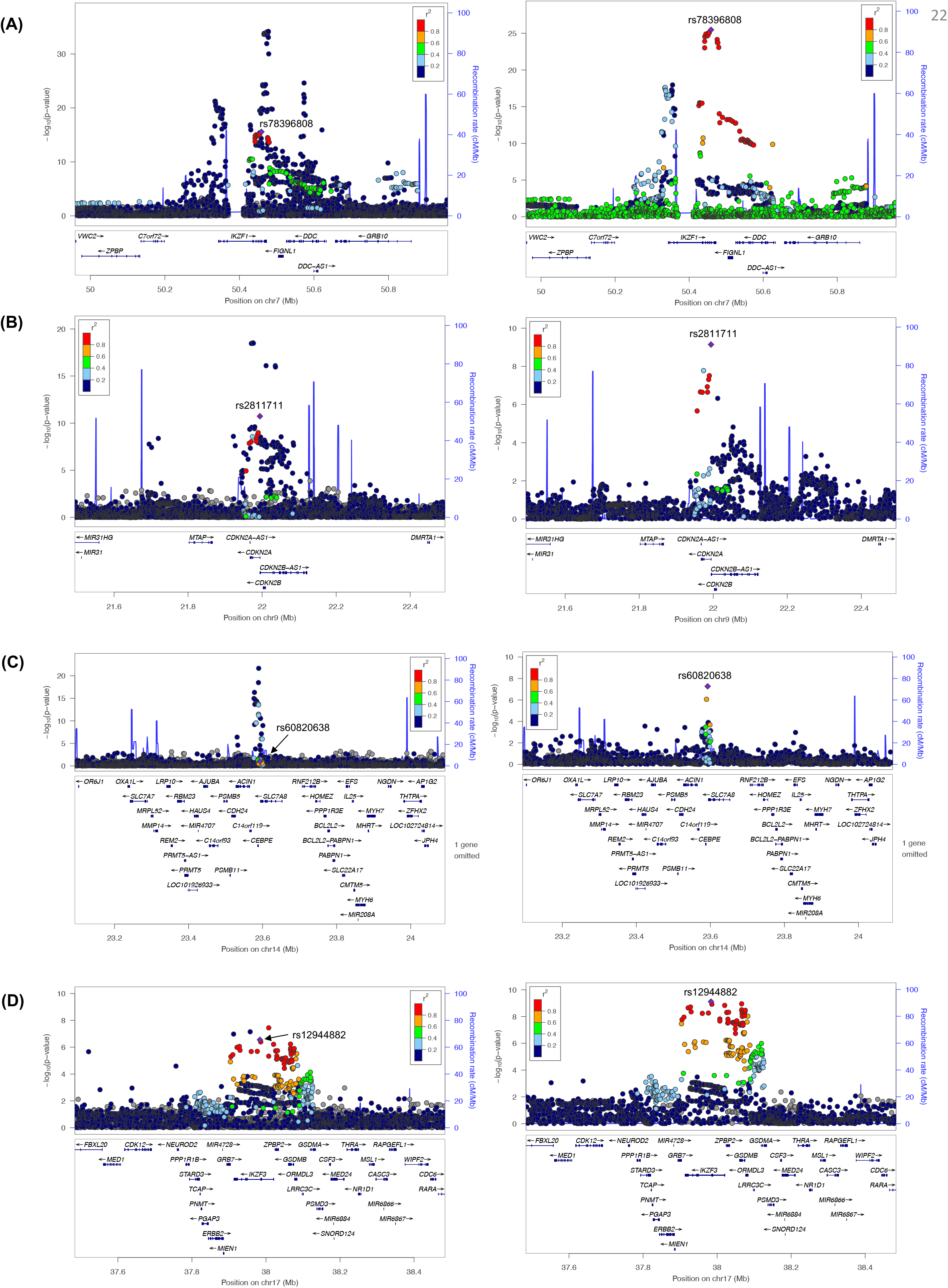
Secondary association signal (p < 5×10^−8^) with ALL found in previously known loci through conditional analysis. LocusZoom plot displaying the 1 Mb region found to harbor a second novel variant associated with ALL through conditional analysis: (A) IKZF1 (B) CDKN2A (C) CEBPE (D) IKZF3. For each locus, we display the pattern of association before(left) and after(right) conditioning on the top associated variant in the locus. In both cases, diamond indicates the lead SNP in the conditional analysis. Color of the remaining SNPs is based on linkage disequilibrium (LD) with the lead variant in the conditional analysis in non-Latino white. Genomic coordinates on x-axis are in hg19.

### Polygenic Risk Score

To assess the combined effect of all identified risk alleles for ALL, we constructed a PRS model in our discovery cohort, using either the 18 SNPs from 16 previously known loci or the 23 known plus novel SNPs and their associated effect sizes from the trans-ethnic meta-analysis. We then computed and tested the PRS for NLW and LAT individuals in the independent CCLS and COG/WTCCC cohorts. The scores generated with the known risk loci were significantly associated with case-control status in all groups (P_CCLS NLW_=2.22×10^−17^, P_CCLS LAT_=4.78×10^−23^, P_COG/WTCCC_ =2.99×10^−62^, Supplementary Table S5). Adding the three novel loci identified in this study and the two novel secondary signals further strengthened the evidence of the association in COG/WTCCC (P =6.93×10^−63^) and CCLS LAT (P = 5.75 ×10^−24^), while the evidence of association stayed about the same in CCLS NLW (P =2.03 ×10^−17^). The predictive accuracy as measured by AUC are similar between NLW and LAT, at around 67-68%, consistent with the hypothesis that trans-ethnic meta-analysis will enable PRS to be more transferrable between populations.

We also examined the distribution of PRS in CCRLP individuals (Figure 4). We found that while the shape of the PRS distribution is consistent with a normal distribution (Kolmogorov-Smirnov P = 0.918 and 0.303 for LAT and NLW, respectively) and appears similar between LAT and NLW (standard deviation of 0.728 and 0.735 respectively; F-Test P = 0.633), the scores in LAT are shifted to the right compared to the scores in NLW (mean of 5.101 and 4.641 respectively, Welch t-test P = 1.3×10^−122^). The observed pattern was consistent when the scores were stratified by case-control status (mean of 5.324 and 4.881 in LAT and NLW cases, respectively, P=3.956 ×10^−58^; mean of 4.895 and 4.414 in LAT and NLW controls, respectively, with P= 1.493 ×10^−78^). This observation was also replicated in CCLS with mean of 5.119 in LAT and 4.607 in NLW (P=4.596 ×10^−51^). Therefore, results from our PRS analyses are consistent with the notion that differences in allele frequency of ALL risk loci between populations may complement other non-genetic factors for ALL risk, and partly explain the increased ALL risk in LAT relative to NLW children and LAT.

**Figure 4.**
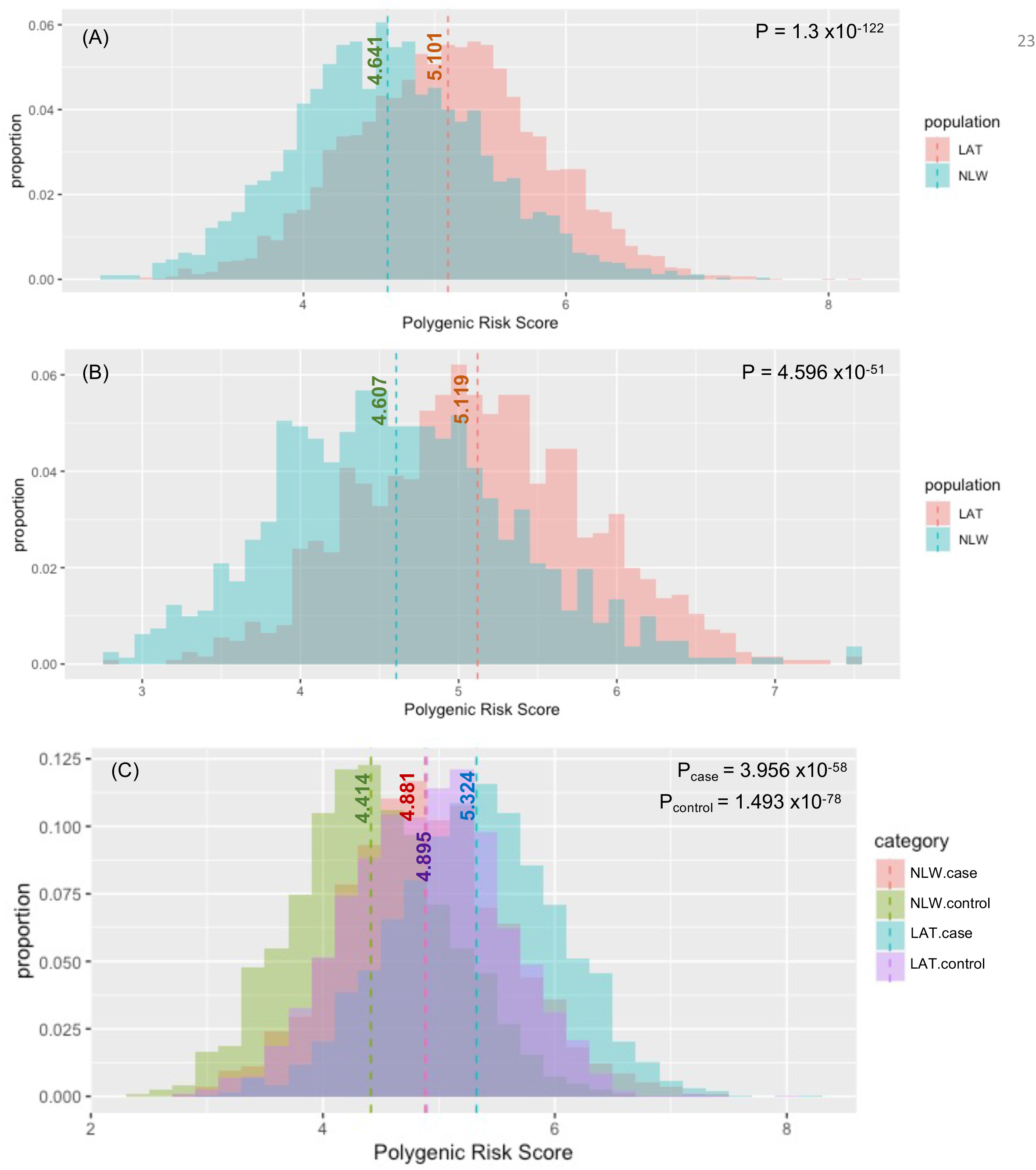
Polygenic Risk Score (PRS) distribution based on GWAS loci for ALL. We compared the PRS distribution between LAT and NLW cohorts in (A) CCRLP and (B) CCLS cohorts. PRS were constructed by summing up imputed dosage weighted by effect size for each Latino(red) and non-Latino white individual(green). In (C) We further stratified the PRS in CCRLP cohort by case/control status. The population mean is indicated with vertical dash lines with the mean score shown. P-values on the right upper corner of each graph is from one-sided t-test comparing the difference in PRS between LAT and NLW overall or within cases and controls.

### Genetic architecture of ALL in Latinos and non-Latino whites

We estimated the relative contributions of each variant to ALL risk by computing the familial relative risk (Supplementary Table 6). In CCLS, where effect size estimates are expected to be less biased by winner’s curse, the known risk variants accounted for 22.7% and 23.2% of familial relative risk in LAT and NLW, and the addition of novel variants increased these estimates to 24.3% and 24.8%, respectively (Supplemental Table S6).

The heritability of ALL attributable to all common SNPs (MAF ≥ 0.05) was estimated to be 20.3 ± 3.2% in NLW and 4.1 ± 2.0% in LAT using the GCTA-LDMS framework^33^, and 20.2 ± 4.7%% in NLW and 11.1 ± 3.6% in LAT using the phenotype-correlation-genotype-correlation (PCGC) regression framework (Supplemental Table S7A). The heritability estimates in NLW are consistent in both approaches and with that previous reported^9(p1)^. Because the imputation quality using HRC reference panel is expected to be high for variants with MAF between 1-5% in NLW, our dataset also provides the opportunity to estimate the frequency-stratified contribution to the heritability of ALL in NLW. The inclusion of low frequency variants increased the estimated heritability in NLW to 29.8 ± 4.3% using REML (divided ∼16.2% due to common variants, 13.5% due to low frequency variants; Supplemental Table S7B). Taking advantage of the admixed nature of LAT, whereby ancestry segments could capture effects beyond that directly attributable to assayed SNPs (such as the estimate from GCTA-LDMS), we also adopted an approach described in Zaitlen et al^34^ to estimate the total narrow-sense heritability for ALL in LAT to be 37.3 ± 6.9%. Taken together, multiple lines of evidence suggest that increasing sample sizes will identify additional low frequency associations to ALL in the future.

Furthermore, we estimated the genetic correlation of ALL between NLW and LAT to be high (rG = 0.714 ± standard error 0.130) but significantly different from 1 (P = 0.014, Supplementary Table 8). This indicates the genetic architectures of NLW and LAT may be similar as expected from correlated effect sizes (Supplementary Figure S1) but not perfectly concordant. We complemented this analysis further by estimating the number of population-specific and shared causal alleles using the program PESCA^35^. The PESCA framework defines the set of causal variants as all variants tested to have a non-zero effect, even if the effect is indirect and only statistical rather than biological in nature. Using this framework, we estimated that approximately 32.5% of SNPs inferred to be causal are shared between NLW and LAT (1.71% of all common SNPs were inferred to have nonzero effects in both NLW and LAT; 1.69% and 1.87% were inferred to have population-specific nonzero effects in NLW and LAT, respectively). Together, these results suggests that there may be ethnic-specific genetic risk profiles or differential interactions with the environment that contributes to differences in disease risk between NLW and LAT. However, it should be noted that these analyses adopted the REML framework or used the GCTA-LDMS estimates as hyperparameters, which could be biased in the context of LAT population here (see Discussion).

## Discussion

By incorporating data across four ethnic groups, we have performed the largest trans-ethnic meta-analysis GWAS of childhood ALL to date. We identified three putatively novel susceptibility loci and two additional independent risk associations at previously reported loci. Our analysis suggests that the known and novel ALL risk alleles together explained about 25% of the familial relative risk in both NLW and LAT populations, and that the trans-ethnic PRS we constructed, although relatively simple and utilizing only the genome-wide associated variants, performed similarly in both NLW and LAT in predicting ALL (AUC ∼ 67-68%).

In support of their potential role in ALL etiology, each of the three novel loci harbors genes and/or variants with a role in hematopoiesis and leukemogenesis as annotated by HaploReg (version 4.1) ^36^ and GTEx portal^37^. The associated variants in 6q23 are located between *HBS1L* and *MYB*, a myeloblastosis oncogene that encodes a critical regulator protein of lymphocyte differentiation and hematopoiesis^38^. This locus is already well known for associations with multiple blood cell measurements, severity of major hemoglobin disorders, and β-thalassemia^39,40^. The associated SNPs in our study fall within *HBS1L-MYB* intergenic region known to harbor multiple variants that reduce transcription factor binding, affect long-range interaction with *MYB*, and impact *MYB* expression^39,41^. The lead SNP rs9376090 is in a predicted enhancer region in K562 leukemia cells and GM12878 lymphoblastoid cells, and is a known GWAS hit for platelet count^38^ and hemoglobin concentration^42,43^. Also, it is an eQTL in lymphocytes and whole blood^37^ for *ALDH8A1*, which encodes aldehyde dehydrogenases, a cancer stem cell marker and a regulator self-renewal, expansion, and differentiation.

One of the associated loci in 10q21 has a distinct haplotype structure, with 130 highly correlated SNPs (r^2^ > 0.8) associated with ALL (Figure 2B). This haplotype structure is observed in LAT and EAS, and the associations are driven by alleles with higher frequency in LAT and EAS than NLW or AFR (Supplementary Table S1, Supplementary Figure S2). This 400kb region is rich with genetic variants associated with blood cell traits such as platelet count, myeloid white cell count, and neutrophil percentage of white cells^44,45^. It is also associated with IL-10 levels^46^ which was shown to be in deficit in ALL cases^47^. The signal region is contained within the intron of *JMJD1C*, a histone demethylase that a recent study has found to regulate abnormal metabolic processes in AML^48^. Previous studies have found that it acts as a coactivator for key transcription factors to ensure survival of AML cells^49^ and self-renewal of mouse embryonic stem cells^50^.

The second locus in 10q21 contains intronic variants in the *TET1* gene, which is well known for its oncogenicity in several malignancies including AML^51^. A recent study showed the epigenetic regulator *TET1* is highly expressed in T-cell ALL and is crucial for human T-ALL cell growth in vivo^52^. We found the associations at this locus to be slightly stronger for T-ALL than for B-ALL in a small subset of individuals with ALL subtype information, though the difference is not statistically significant (Supplemental Table S9). Of the four significant variants in this locus, SNP rs58627364 lies in the promoter region of *TET1* while the remaining three variants did not appear to overlap functional elements (Supplementary Figure S3). However, none of these SNPs were observed as eQTL for *TET1* in whole blood or lymphoblastoid cells^53^; future studies may want to investigate whether these SNPs affect *TET1* expression in hematopoietic stem or progenitor cells.

In addition to identifying putative novel ALL risk loci, we capitalized on the large numbers of Latinos and non-Latino whites included in our study to explore the genetic architecture of ALL in these two populations. In the NLW population, we estimated that ∼ 29% of the heritability of ALL was attributed to a combination of common and low-frequency (MAF between 1-5%) imputed variants using both GCTA-LDMS and PCGC regression (Supplementary Table S7B). This estimate is higher than the previous estimate of ∼20% (ref. ^9^), suggesting that there are additional low frequency variants associated with ALL that may be discovered in larger scaled studies. The picture is less clear among LAT, where the estimated heritability was perhaps unrealistically low using GCTA-LDMS (4.1% in univariate analysis; Supplementary Table S7A, 8). This estimate contrasted strongly against other lines of evidence that showed similar estimated effect sizes (r2 = 0.819; Supplementary Figure S1) and familial relative risks explained by GWAS loci (Supplementary Table S6) between NLW and LAT. Previous studies have noted the downward bias in REML heritability estimate in case-control studies, which is exacerbated when the covariates in the model (*i*.*e*. PCs and ancestry) are correlated with the disease status^54^. We thus also followed previous suggestions and used the PCGC regression to obtain variance component estimates^54,55^, resulting in a higher heritability estimate (11.1%). Because of the underrepresentation of Native American or other non-European haplotypes in HRC panel, *a priori* we did not estimate the heritability including low-frequency imputed variants in LAT. When we did attempt to estimate heritability in this setting, we obtained a strongly negative REML-based heritability estimates (Supplementary Table S7B), suggesting potential model instability or misspecification attributed to the admixed nature of LAT^56^. Consequently, we also recommend caution when interpreting the estimated genetic correlation between LAT and NLW.

Nevertheless, we used bivariate version of the REML analysis to compute genetic correlation of ALL between populations, as had been done previously for prostate cancer with individual level data^57^. Our estimated genetic correlation (r_G_ = 0.71) is significantly less than 1, apparently suggesting a significant population-specific components of the disease architecture between LAT and NLW. This would be consistent with the findings of the *ERG* locus^16,17^, a Latino-specific association with ALL, and suggest that future ethnic-specific GWAS across different ethnic groups for ALL will be insightful. This is also consistent with our observation in the PESCA analysis, where we found that only 32.5% of the estimated causal alleles are shared between LAT and NLW. These insights should still be treated with caution because the sample size for ALL, a rare disease, is still relatively small compared to complex traits examined using PESCA^35^, and because the REML-based heritability estimates for LAT used as hyperparameter by PESCA may be biased. Therefore, more focused efforts to investigate the genetic architecture for ALL, particularly in admixed populations like the Latinos, is needed.

Future studies aimed to uncover the genetic risk factors for ALL could focus on multiple avenues. First, there will be a need to further increase the sample size of the study cohort, which would provide additional venues to replicate the putative novel findings here and identify more associated alleles at lower frequency. Second, there should be a focus on ethnic-specific GWAS for ALL, as ethnic-specific associations could be missed in a trans-ethnic GWAS. An example is the *ERG* locus, which is not genome-wide significant in our meta-analysis. Finally, while not explored extensively in this particular study, there should be a focus on disentangling the different subtypes of ALL, and to study other aspects of the disease pathogenesis such as disease progression or risk of relapse, though these data are less available and may require more focused ascertainment and cohort creation.

## Data Availability

Any uploading of genomic data and/or sharing of these biospecimens or individual data derived from these biospecimens has been determined to violate the statutory scheme of the California Health and Safety Code Sections 124980(j), 124991(b), (g), (h), and 103850 (a) and (d), which protect the confidential nature of biospecimens and individual data derived from biospecimens. This study was approved by Institutional Review Boards at the California Health and Human Services Agency, University of Southern California, Yale University, and the University of California San Francisco. The de-identified newborn dried blood spots for the CCRLP (California Biobank Program SIS request # 26) were obtained with a waiver of consent from the Committee for the Protection of Human Subjects of the State of California.

## Acknowledgement

This work was supported by research grants from the National Institutes of Health (R01CA155461, R01CA175737, R01ES009137, P42ES004705, P01ES018172, P42ES0470518 and R24ES028524) and the Environmental Protection Agency (RD83451101), United States. The content is solely the responsibility of the authors and does not necessarily represent the official views of the National Institutes of Health and the EPA. The collection of cancer incidence data used in this study was supported by the California Department of Public Health as part of the statewide cancer reporting program mandated by California Health and Safety Code Section 103885; the National Cancer Institute’s Surveillance, Epidemiology and End Results Program under contract HHSN261201000140C awarded to the Cancer Prevention Institute of California, contract HHSN261201000035C awarded to the University of Southern California, and contract HHSN261201000034C awarded to the Public Health Institute; and the Centers for Disease Control and Prevention’s National Program of Cancer Registries, under agreement U58DP003862-01 awarded to the California Department of Public Health. The biospecimens and/or data used in this study were obtained from the California Biobank Program, (SIS request #26), Section 6555(b), 17 CCR. The California Department of Public Health is not responsible for the results or conclusions drawn by the authors of this publication. We thank Hong Quach and Diana Quach for DNA isolation support. We thank Martin Kharrazi, Robin Cooley, and Steve Graham of the California Department of Public Health for advice and logistical support. We thank Eunice Wan, Simon Wong, and Pui Yan Kwok at the UCSF Institute of Human Genetics Core for genotyping support. This study makes use of data generated by the Wellcome Trust Case–Control Consortium. A full list of the investigators who contributed to the generation of the data is available from www.wtccc.org.uk. Funding for the project was provided by the Wellcome Trust under award 076113 and 085475. Genotype data for COG ALL cases are available for download from dbGaP (Study Accession: phs000638.v1.p1). Data came from a grant, the Resource for Genetic Epidemiology Research in Adult Health and Aging (RC2 AG033067; Schaefer and Risch, PIs) awarded to the Kaiser Permanente Research Program on Genes, Environment, and Health (RPGEH) and the UCSF Institute for Human Genetics. The RPGEH was supported by grants from the Robert Wood Johnson Foundation, the Wayne and Gladys Valley Foundation, the Ellison Medical Foundation, Kaiser Permanente Northern California, and the Kaiser Permanente National and Northern California Community Benefit Programs. The RPGEH and the Resource for Genetic Epidemiology Research in Adult Health and Aging are described here: https://divisionofresearch.kaiserpermanente.org/genetics/rpgeh/rpgehhome. For recruitment of subjects enrolled in the CCLS replication set, the authors gratefully acknowledge the clinical investigators at the following collaborating hospitals: University of California Davis Medical Center (Dr. Jonathan Ducore), University of California San Francisco (Drs. Mignon Loh and Katherine Matthay), Children’s Hospital of Central California (Dr. Vonda Crouse), Lucile Packard Children’s Hospital (Dr. Gary Dahl), Children’s Hospital Oakland (Dr. James Feusner), Kaiser Permanente Roseville (formerly Sacramento) (Drs. Kent Jolly and Vincent Kiley), Kaiser Permanente Santa Clara (Drs. Carolyn Russo, Alan Wong, and Denah Taggart), Kaiser Permanente San Francisco (Dr. Kenneth Leung), and Kaiser Permanente Oakland (Drs. Daniel Kronish and Stacy Month). The authors additionally thank the families for their participation in the California Childhood Leukemia Study (formerly known as the Northern California Childhood Leukemia Study). Finally, the authors acknowledge the Center for Advanced Research Computing (CARC; https://carc.usc.edu) at the University of Southern California for providing computing resources that have contributed to the research results reported within this publication.

## Authorship

### Contribution

J.L.W., C.W.K.C., A.J.D. conceived and supervised this project; S.J., S.L., M.C. performed data analysis; N.M., I.S.M., L.M.M., A.T.D., C.M., X.M. provided resources; S.J., A.J.D., C.W.K.C., J.L.W. wrote the manuscript with input from all the coauthors.

### Correspondence

Joseph Leo Wiemels, Center for Genetic Epidemiology, 1450 Biggy St, Los Angeles, California; Charleston Chiang, 1450 Biggy St, Los Angeles, California

---

**Figure S1.**
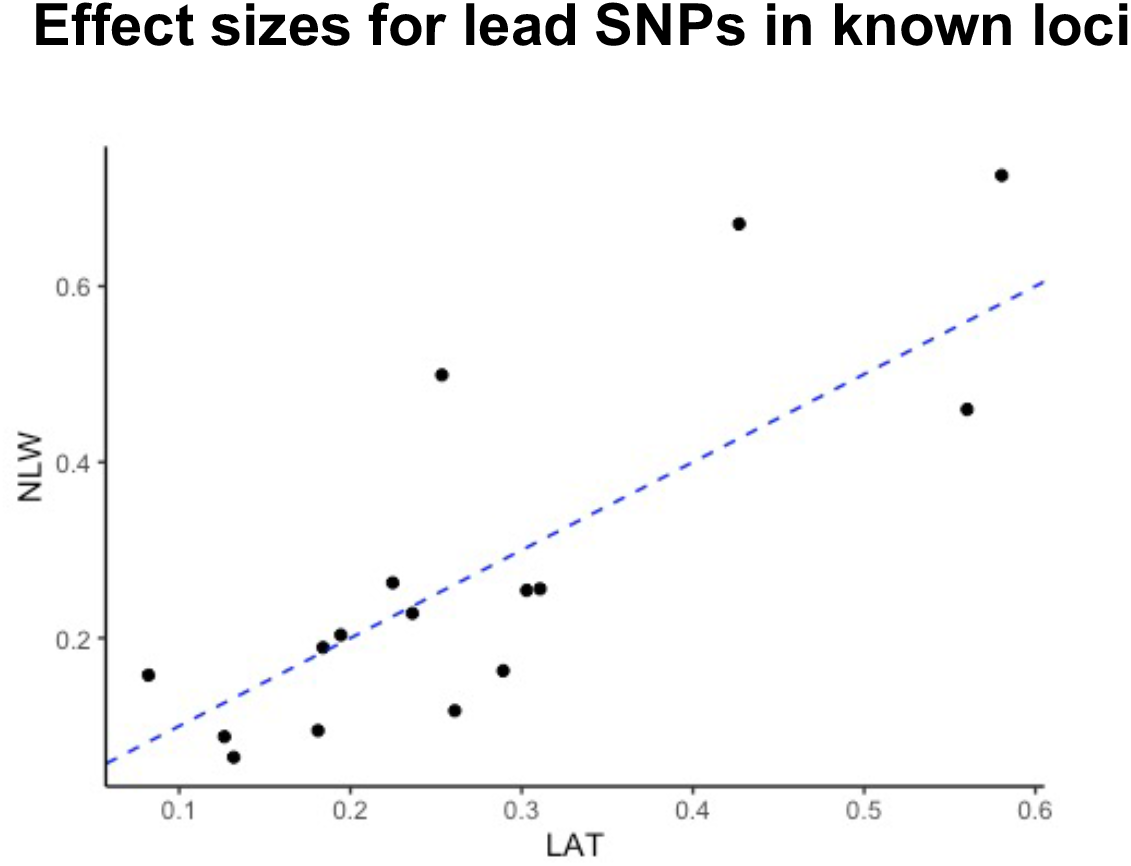
Effect sizes for lead SNPs in each of the 16 known loci. The effect size estimates(Beta) from GWAS in only the LAT or NLW subset from our discovery cohort are shown. The correlation coefficient (r) is 0.819. The dashed blue line is y=x.

**Figure S2.**
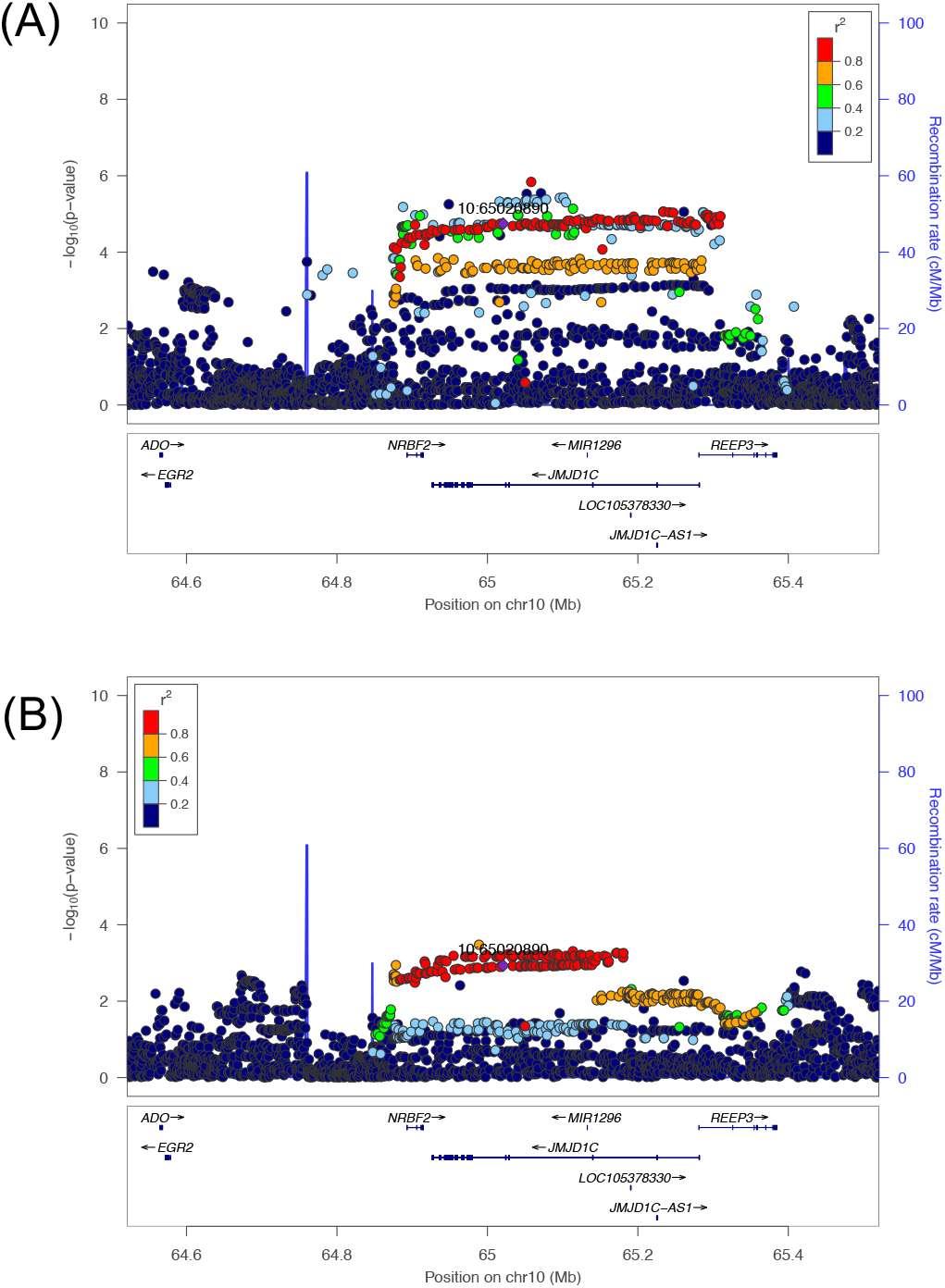
Association signal around NRBF2/JMJD1C locus on chr10 in LAT and EAS cohorts. LocusZoom plots show distinct haplotypes showing association with ALL in (A) LAT and (B) EAS cohorts in our study. Diamond symbol indicates the lead SNP in each cohort. Color of remaining SNPs is based on linkage disequilibrium (LD) as measured by r2 with the lead SNP in the respective cohort. All coordinates in x-axis are in hg19.

**Figure S3.**
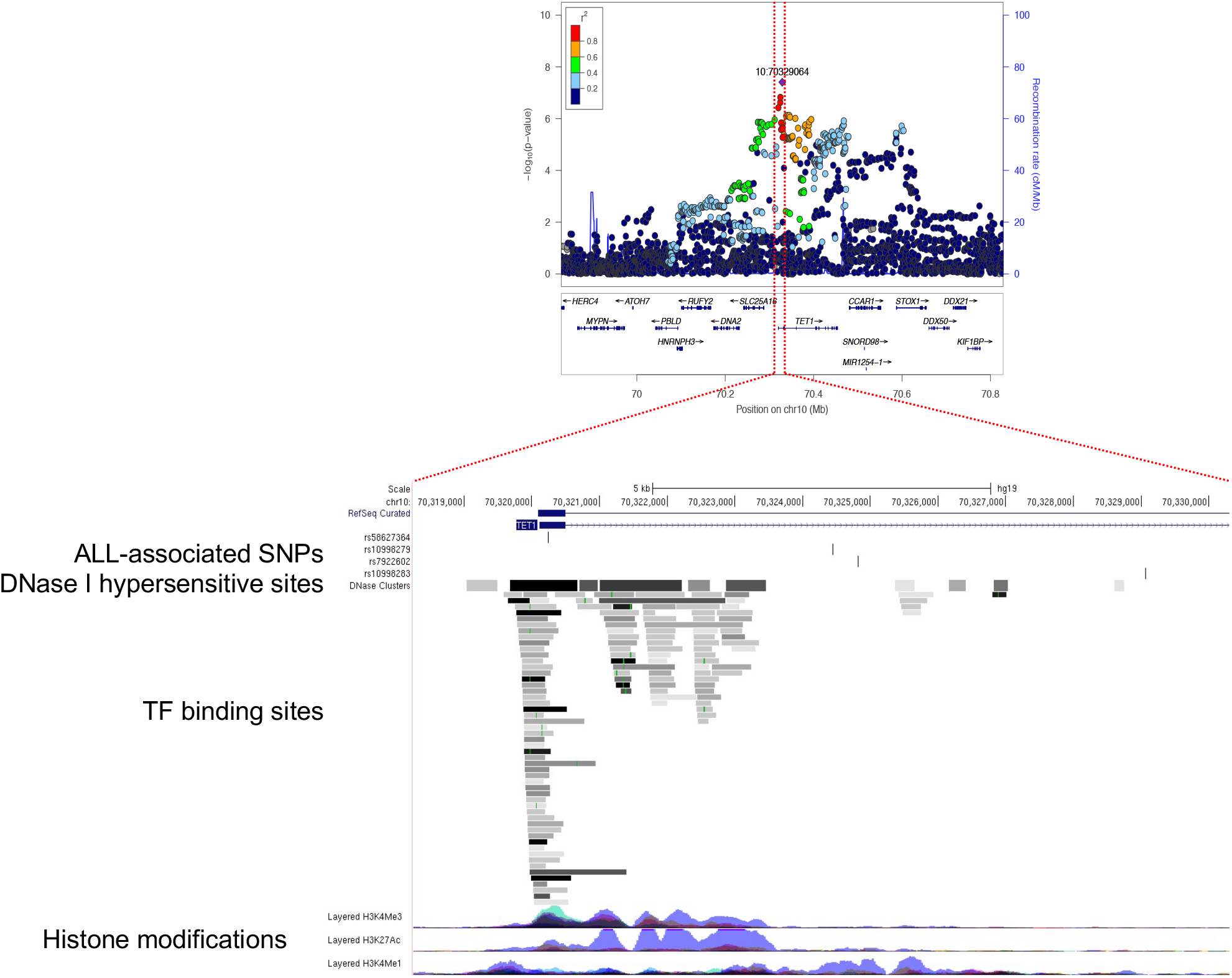
Functional annotation of the TET1 locus. For the immediately nearby location around the top associated SNPs in our meta-analysis(blue vertical lines), we extracted the functionally annotated genomic/epigenomic features from multiple cell types in ENCODE data. Functional data were retrieved from UCSC genome browser.

