## Supplemental Figures for "Genome-wide trans-ethnic meta-analysis identifies novel susceptibility loci for childhood acute lymphoblastic leukemia"

### Slide 1
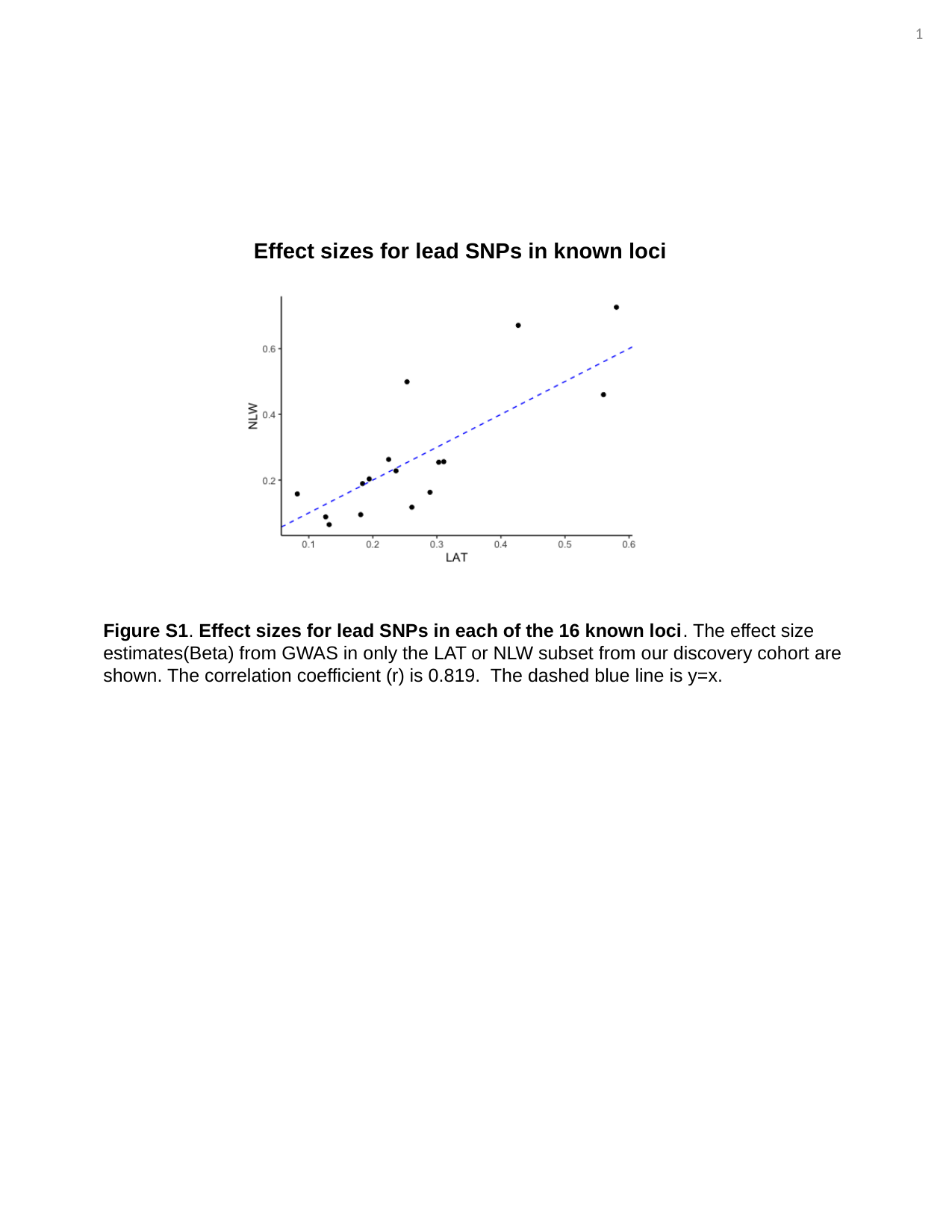

19
Effect sizes for lead SNPs in known loci
Figure S1. Effect sizes for lead SNPs in each of the 16 known loci. The effect size estimates(Beta) from GWAS in only the LAT or NLW subset from our discovery cohort are shown. The correlation coefficient (r) is 0.819. The dashed blue line is y=x.

### Slide 2
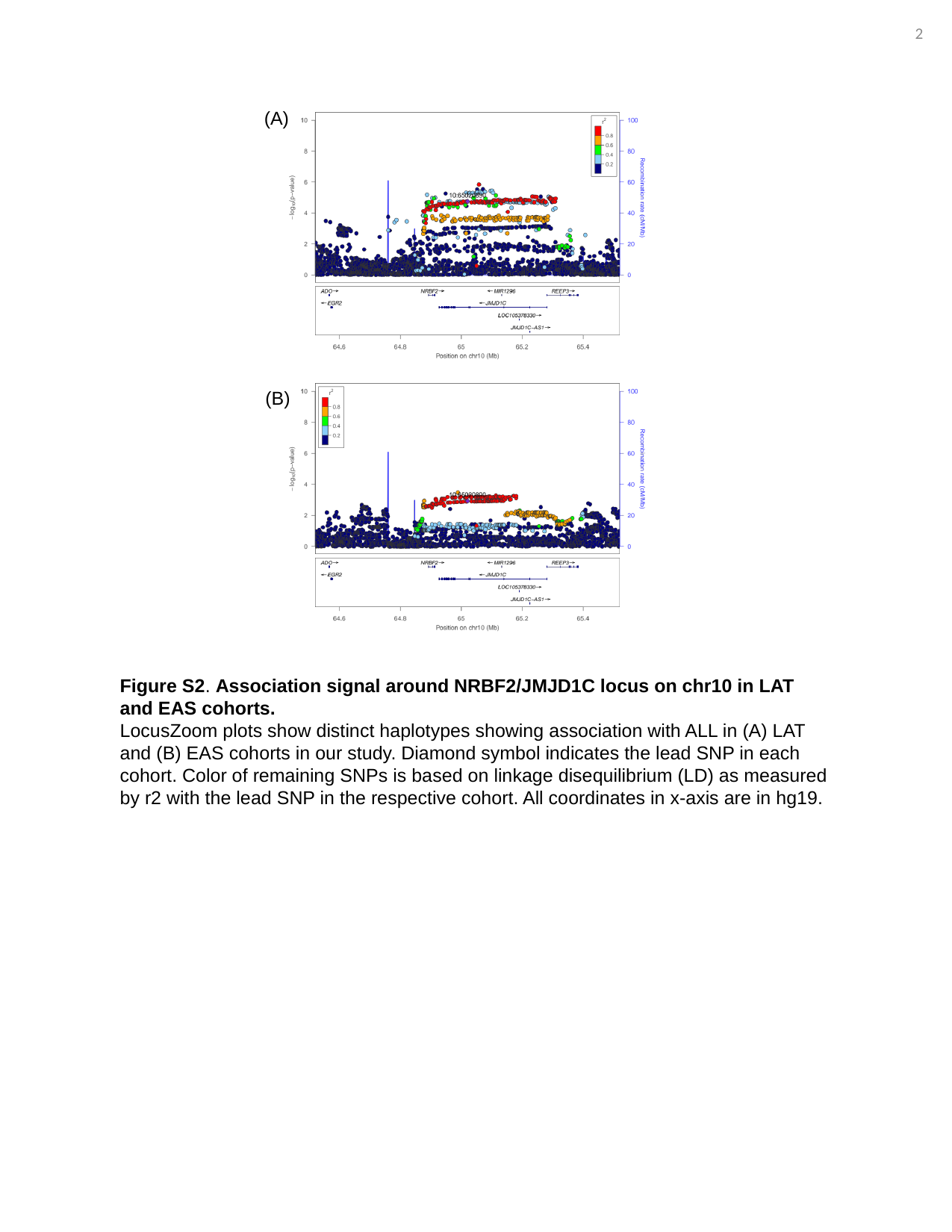

20
(A)
(B)
Figure S2. Association signal around NRBF2/JMJD1C locus on chr10 in LAT and EAS cohorts.
LocusZoom plots show distinct haplotypes showing association with ALL in (A) LAT and (B) EAS cohorts in our study. Diamond symbol indicates the lead SNP in each cohort. Color of remaining SNPs is based on linkage disequilibrium (LD) as measured by r2 with the lead SNP in the respective cohort. All coordinates in x-axis are in hg19.

### Slide 3
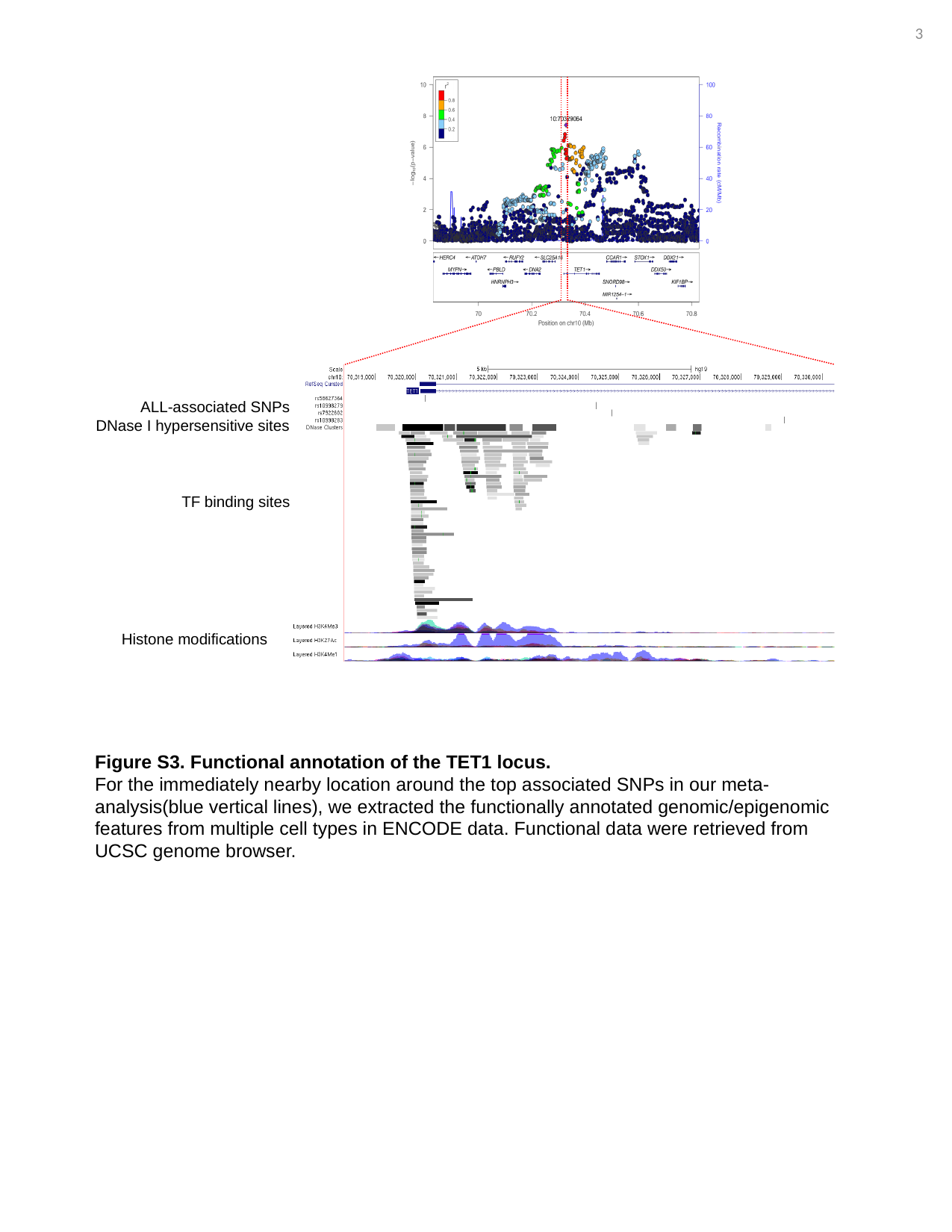

21
ALL-associated SNPs
DNase I hypersensitive sites
TF binding sites
Histone modifications
Figure S3. Functional annotation of the TET1 locus.
For the immediately nearby location around the top associated SNPs in our meta-analysis(blue vertical lines), we extracted the functionally annotated genomic/epigenomic features from multiple cell types in ENCODE data. Functional data were retrieved from UCSC genome browser.
