## Supplemental Methods for "Genome-wide trans-ethnic meta-analysis identifies novel susceptibility loci for childhood acute lymphoblastic leukemia"

**Data Processing and Quality Control**

The quality control (QC) on single nucleotide polymorphism (SNP) array genotypes and samples were carried out in each population and dataset in parallel, performed in two stages: pre-imputation and post-imputation. In pre-imputation QC, the sex chromosomes were excluded, and SNPs were filtered out on the basis of call rate (<98%), minor allele frequency (MAF<0.01), genome-wide relatedness (PI_HAT>0.02), genome heterozygosity rate (mean heterozygosity± 6Std), and deviation from Hardy-Weinberg equilibrium in controls (P<10^-5^). Samples with call rate < 95% were also removed. In general, individuals were included in each of the four ethnic groups based on their self-reported ethnicity. We did not attempt to reassign individuals to different ethnic groups based on estimated genetic ancestry. We performed principal components analysis for each population including our study subjects along with 1000 Genomes Project reference data^1^ to identify extreme outlier individuals that clustered apart from other individuals in their self-reported race/ethnicity groups: this identified a small subset of individuals (29 cases and 51 controls from CCRLP, 31 individuals from GERA) among self-reported Asians (total n=722) that clustered with South Asian reference individuals, as well as 5 self-reported African American individuals out of n=3572 from the GERA cohort that clustered with East Asian reference individuals. These appear to result from a lack of finer-scale ethnic labels for self-report, or a mis-labeling of individual records, and these individuals were dropped from our analysis.

To control for potential batch effect and systematic bias between array types, we further performed two separate GWASs for quality control purposes. First, stratified by ethnicity and restricted to SNPs passing the filters described above in both CCRLP and GERA, we compared CCRLP controls and GERA individuals. Second, using the GERA NLW cohort we compared individuals that were genotyped on the Axiom type “A” to those genotyped on the type “O” reagent kit (NLW was the only cohort in GERA that was genotyped using both reagent kits). Twenty principal components (PCs) were included as covariates of the logistic regression. In both comparisons we observed inflation of the test statistics suggesting a subset of SNPs exhibited evidence of batch effect, thus we removed variants with P < 0.01 in any of the comparisons from all populations.

We then performed genome-wide imputation with the overlapping set of remaining SNPs (N = 431,543 in AFR, 259,468 in EAS, 547,575 in LAT and 362,977 in NLW) in each dataset using Haplotype Reference Consortium (HRC v r1.1 2016) as a reference in the Michigan Imputation Server^2^. The different number of SNPs passing QC and used in imputation reflects the fact that each ethnic group in CCRLP was genotyped using Affymetrix World arrays optimized for the Latino population (i.e., Axiom LAT array). In post-imputation QC, we filtered variants in each ethnic group by imputation quality (R^2^ < 0.3), MAF (< 0.01), and allele frequency difference between non-Finnish Europeans in the Genome Aggregation Database (gnomAD)^3^ and CCRLP NLW controls ( > 0.1). We next performed another GWAS between CCRLP controls and GERA individuals to conservatively protect against between-cohort batch effects after imputation, and removed variants with P < 1x10^-5^. In principal components analysis using imputed data, we identified and removed 31 individuals in GERA LAT that were extreme outliers after imputation in PCs 1 to 20. Stratified by ethnicity, the CCRLP and GERA datasets were then merged to perform GWAS of ALL. In total, 124, 318, 1878, 1162 cases and 2067, 5017, 8410, 57341 controls, in AFR, EAS, LAT and NLW, respectively were used in GWAS for ALL. An effective population was calculated using this equation:

$$N_{eff}= \frac{4}{\frac{1}{N_{cases}}+\frac{1}{N_{controls}}}$$

where *N_eff_* is the effective sample size, *N_cases_* is the number of cases, and *N_controls_* is the number of control subjects. A total of 7,628,894 SNPs that remained in at least three ethnic groups were tested in our GWAS discovery analysis.

For replication cohorts, we generally followed the same quality control pipeline. For COG and WTCCC, because self-identified ethnicity was not available to us, we performed global ancestry estimations using ADMIXTURE and the 1000 Genomes populations as reference and removed individuals with < 90% estimated European ancestry from the analysis. This resulted in a total of 1504 and 2931 NLW cases and controls, respectively, from COG/WTCCC, and 426 NLW cases, 278 NLW controls, 758 LAT cases and 549 LAT controls, from CCLS.

**Familial risk per variant**

The percentage of familial relative risk (FRR) explained by each genetic variant was calculated as per Schumacher et al^4^ . The familial relative risk due to locus $k$ ($\lambda_{k}$) is given by

$\lambda_{k}=\frac{p_{k}r_{k}^{2}+q_{k}}{\left( p_{k}r_{k}+q_{k} \right)^{2}}$

where $p_{k}$ is the frequency of the risk allele for locus $k$ in each population, $q_{k}=1-p_{k}$, and $r_{k}$ is the estimated per-allele odds ratio from meta-analysis. The percentage of familial relative risk is calculated as $\sum_{k} \log\lambda_{k}/\log\lambda_{0}$ where $\lambda_{0}$ is the observed familial risk to first-degree relatives of ALL cases, assumed to be 3.2 as per Kharazmi et al^5^.

**Heritability Estimates**

We estimated heritability ascribable to all post-QC imputed SNPs with MAF $\geq$0.05 in our GWAS data using the genome-wide complex trait analysis software (GCTA)^6^. We followed the GCTA-LDMS approach to estimate heritability from imputed data^7^, which recommended stratifying SNPs into bins based on their LD scores and/or minor allele frequency. Using GCTA, we computed the genetic relationship matrix (GRM) of pairs of samples using SNPs in each bin, and used the multiple GRMs as input to obtain a restricted maximum likelihood (REML) estimate of heritability. All individuals in discovery analysis were used for LAT (n=10,288). For computational efficiency and for maintaining a close balance in sample size to the LAT data, we randomly sampled 10,000 NLW GERA controls to be included with all of CCRLP NLW cases and controls (total N = 12,391). We used a prevalence of 4.41x10^-4^ and 4.09x10^-4^ for childhood ALL in LAT and NLW respectively based on data from the Surveillance Research Program, (National Cancer Institute SEER*Stat software version 8.3.8; https://seer.cancer.gov/seerstat) to convert the estimated heritability to the liability scale. Because the NLW are expected to be much better imputed using HRC than LAT, particularly at rare variants, our genome-wide imputed data potentially could be used to partition the contribution of low frequency (0.01 ≤ MAF < 0.05) and common (MAF $\geq$ 0.05) variants in NLW population. In this case, we performed GCTA-LDMS analysis in 8 strata: two MAF strata (low frequency and common) by four quartiles of LD score strata. We also used these same GRMs to estimate heritability using phenotype-correlation-genotype-correlation (PCGC) regression as implemented in LDAKv5.1^8,9^.

We further applied an approach to estimate heritability in LAT population using local ancestry^10^. In brief, we first estimated the local ancestry in LAT using RFMix^11^, using the combined 1000 Genomes Project and Human Genome Diversity Project^3^ as ancestry references. Specifically, we used AFR (excluding ACB and ASW inidividuals; n=716), self-reported Non-Finnish European (NFE; n=617), and subjects having > 85% global AMR ancestry(based on ADMIXTURE^12^; n=94) as the reference for African, European, and Native American ancestries. We used the local ancestry to estimate the genetic similarity and the heritability explained by local ancestry *h^2^*_γ_, calculated the genetic distance *F*_STC_ between the ancestral populations, and the mean admixture proportion *θ*. Following Zaitlen et al^10^, the heritability is then calculated using formula ${h^{2}}_{\gamma}=2 \theta\left( 1-\theta\right) h^{2}F_{STC}$. Because the original approach is only applicable to two-way admixed populations, we assigned the ancestry call as missing if the most likely local ancestry call for a locus is AFR. This sets approximately 5% of the genome as missing. The number of copies of local ancestry are standardized to have zero mean and unit variance to compute the genetic similarity matrix. The heritability explained by local ancestry *h2* was estimated in GCTA, with the global AMR ancestry normalized by non-African ancestry as a quantitative covariate. The genetic distance $F_{STC}$ between the ancestral populations was computed based on the allele frequencies in the reference population as $\frac{{(f_{AMR}-f_{NFE)})}^{2}}{2f (1-f)}$ where $f_{AMR}$ and $f_{NFE}$are allele frequencies in AMR and NFE reference panel and the expected frequency in the admixed population, $f$, is the average of ancestral frequencies weighted by the average normalized global ancestries. The genome wide *F_STC_* is the average value across 417,635 sites where the minor allele frequencies are greater than 0.05 in both ancestral populations.

To measure genetic correlation between LAT and NLW, we used SNPs with MAF $\geq$ 0.05 in both populations to generate GRM using R as per Mancuso et al.^13^ The individuals used in univariate REML for each ethnicity were used for the bivariate analysis (n=22,679). We used imputed dosage data to estimate GRM for each unique pair of ancestry groups as

$$A=\frac{1}{m}\left[ \begin{matrix} Z_{1}Z_{1}^{t} & Z_{1}Z_{2}^{t} \\ Z_{2}Z_{1}^{t} & Z_{2}Z_{2}^{t} \end{matrix} \right]$$

where m is number of SNPs and Z_1_ and Z_2_ are the standardized genotype matrices for LAT and NLW, respectively. We estimated genetic correlation using bivariate GREML in GCTA^6^.

**Investigation of Genetic Architecture**

To quantify the extent to which latent causal variants for ALL are shared or population-specific between LAT and NLW, we analyzed our GWAS summary data using the tool PESCA^14^. Briefly, PESCA analyzes GWAS summary data from multiple populations jointly to infer the genome-wide proportion of causal variants that are population-specific or population-shared. For computational efficiency, PESCA requires first defining LD blocks that are approximately independent in both populations and assumes that a SNP in a given block is independent from all SNPs in all other blocks. We computed pairwise LD matrix in both NLW and LAT using ~329K directly genotyped SNPs shared in both populations. Then, following Shi et al.^14^, we generated the trans-ethnic LD matrix by using the larger r^2^ value of the NLW or LAT-specific pairwise LD, and used LDetect ^15^ to define LD blocks within the transethnic LD matrix. By setting mean LD block size to 200 SNPs and using default parameters, we obtained 1,653 blocks that are approximately independent, which is approximately similar to previous reports in East Asians and Europeans.^14^ We then followed Shi et al. to estimate the numbers of population-specific and shared causal SNPs using PESCA^14^. We restricted our analysis to 1.3M SNPs with MAF > 0.05, r^2^ < 0.95, and with summary association statistics available in both NLW and LAT. We first estimated the genome-wide proportion of population-specific and shared causal variants with the heritability estimated above (0.2033 and 0.0413 in NLW and LAT, respectively) using default parameters in PESCA, parallelizing the analysis in groups of 10 LD blocks at a time. Using the estimated genome-wide proportions of population-specific and shared causal variants as prior probabilities, we then estimated the posterior probability of each SNP to be causal in a single population (population-specific) or both populations (shared), and inferred the posterior expected numbers of population-specific/shared causal SNPs in each LD block by summing the per-SNP posterior probabilities of being causal in a single or both populations. Critically, while PESCA is an analysis based on summary statistics and not designed for admixed populations, it can be applied to admixed population such as LAT if in-sample LD is used.^14^
